# Mathematical assessment of the role of waning and boosting immunity against the BA.1 Omicron variant in the United States

**DOI:** 10.1101/2022.07.21.22277903

**Authors:** Salman Safdar, Calistus N. Ngonghala, Abba B. Gumel

## Abstract

Three safe and effective vaccines against SARS-CoV-2 (the Pfizer-BioNTech, Moderna and Johnson & Johnson vaccines) have played a major role in combating the COVID-19 pandemic in the United States. However, the effectiveness of these vaccines and vaccination programs has been challenged by the emergence of new SARS-CoV-2 variants of concern. A new mathematical model is formulated to assess the population-level impact of the waning and boosting of vaccine-derived and natural immunity against the Omicron variant in the United States. To account for gradual waning of vaccine-derived immunity, we considered three vaccination classes (*V*_1_, *V*_2_ and *V*_3_; where subscripts 1, 2 and 3 represent high, moderate and low levels of immunity, respectively). The disease-free equilibrium of the model was shown to be globally-asymptotically stable, for two special cases, whenever a certain associated epidemiological quantity, known as the *vaccination reproduction number* of the model, is less than one. The model was fitted using observed daily case data for the Omicron BA.1 variant in the United States. Simulations of the resulting parameterized model showed that, for the case where the high-level of the vaccine-derived protective efficacy received by individuals in the first vaccinated class (*V*_1_) is set at its baseline value (85%; while the vaccine-protective efficacy for individuals in the *V*_2_ and *V*_3_ classes, as well as natural immunity, are maintained at baseline), population-level herd immunity can be achieved in the United States *via* vaccination-boosting strategy, if at least 59% of the susceptible populace is fully-vaccinated followed by the boosting of about 71.5% of the fully-vaccinated individuals whose vaccine-derived immunity has waned to moderate or low level. However, if the high level of vaccine-induced efficacy for individuals in the *V*_1_ class is reduced to 55%, for instance, achieving herd immunity requires fully-vaccinating at least 91% of the susceptible population (followed by marginal boosting of those in whom the vaccine-derived immunity has waned to moderate or low level). In the absence of boosting of vaccine-derived and natural immunity, waning of immunity (both vaccine-derived and natural) only causes a marginal increase in the average number of new cases at the peak of the pandemic. Boosting of both immunity types at baseline could result in a dramatic reduction in the average number of daily new cases at the peak, in comparison to the corresponding waning scenario without boosting of immunity. Furthermore, boosting of vaccine-derived immunity (at baseline) is more beneficial (in reducing the burden of the pandemic) than boosting of natural immunity (at baseline). Specifically, for the fast waning of immunity scenario (where both vaccine-derived and natural immunity are assumed to wane within three months), boosting vaccine-derived immunity at baseline reduces the average number of daily cases at the peak by 90% (in comparison to the corresponding scenario without boosting of the vaccine-derived immunity), whereas boosting of natural immunity (at baseline) only reduced the corresponding peak daily cases (in comparison to the corresponding scenario without boosting of natural immunity) by 62%. It was further shown that boosting of vaccine-derived (implemented near the baseline level) increased the prospects of altering the trajectory of COVID-19 from persistence to possible elimination (even for the fast waning scenario of vaccine-derived immunity). Thus, a vaccination strategy that emphasizes boosting of immunity would significantly enhance the prospects of SARS-CoV-2 elimination in the United States.

## 1. Introduction

Since December 2019, the world has been experiencing a devastating pandemic of a novel coronavirus (COVID-19), caused by SARS-CoV-2, on a scale never before seen since the 1918/1919 influenza pandemic [1]. As of mid July 2022, the SARS-CoV-2 pandemic has caused over 555 million confirmed cases and over 6.35 million deaths globally [2, 3] (with the United States bearing the brunt of the burden, with over 88.6 million confirmed cases and over 1 million COVID-19 deaths) [3]. For most parts of the year 2020, the control and mitigation efforts against SARS-CoV-2 in the United States were restricted to the use of nonpharmaceutical interventions, such as social-distancing, quarantine of suspected cases, isolation of those with symptoms of SARS-CoV-2, use face coverings, community lockdowns, contact-tracing, etc. [4–8], until the Food and Drug Administration (FDA) provided Emergency Use Authorization (EUA) to two safe and highly-efficacious vaccines (developed by Pfizer Inc. and Moderna Inc.) in December of 2020 [9, 10]. Both of the approved vaccines were primarily administered in two-dose regiments with three to four weeks apart, and each offer an estimated protective efficacy against symptomatic COVID-19 infection of about 95% [11, 12]. Another vaccine, developed by Johnson & Johnson (administered as a single dose), received FDA-EUA in late February 2021 [13] (this vaccine has an estimated 75% efficacy in preventing severe/critical illness caused by COVID-19 [14]). The rapid development and administrative deployment of effective vaccines has played an extremely vital role in minimizing and mitigating the global burden of the pandemic [15, 16]. Our study is focused on these three vaccines being used and administered in the United States.

Despite the rapid development and deployment of the effective vaccines, COVID-19 cases and mortality continued to rise in the United States for most part of 2021 (and even early 2022). This is largely due to emergence of deadly and highly-contagious SARS-CoV-2 variants of concern (notably the Alpha, Beta, Gamma, Delta and Omicron variants) [17–21]. Specifically, the emergence of the Omicron variant (B.1.1.529), in November of 2021, has dramatically changed the trajectory of the pandemic [22]. It is believed to be at least three times more contagious than Delta [22, 23]. A subvariant of Omicron, BA.2 was first identified in the United States from a sample collected on December 14, 2021, in New Jersey [24]. It is believed to be more contagious than Omicron i.e. BA.1 [25, 26].

Numerous clinical studies have shown that the efficacy of the SARS-CoV-2 vaccines wane over time (with estimated waning time of about 9 months) [8, 27]. Consequently, the FDA approved the administration of booster shots, for all three vaccines, during August-November of 2021. Primarily, a booster dose (for persons aged 18 years and above), were approved because of waning vaccine effectiveness over time [28]. In late March 2022, the FDA authorized a second booster shot of COVID-19 vaccines for vulnerable populations in the U.S. (i.e., for people 50 years of age and older, and for individuals with certain immuno-compromising conditions who are at higher risk of severe disease, hospitalization and death). A second booster shot is equivalent to a fourth dose for people who received a Pfizer-BioNTech or Moderna mRNA series or a third dose for those who received the single-shot Johnson & Johnson vaccine. The goal of this study is to use mathematical modeling and analysis to assess the population-level impact of vaccination and booster shots (keeping in mind the waning efficacies of all the approved vaccinations) programs, based on using the three FDA-approved vaccines, on the dynamics of the Omicron SARS-CoV-2 variant in the United States. The focus of the study is on determining the minimum vaccination coverage (i.e., vaccine-derived *herd immunity*) needed to effectively curtail the spread of the highly-contagious Omicron variant in the United States.

Numerous mathematical models, of various types, have been formulated and used to gain insight and understanding on the dynamics of the COVID-19 pandemic (with majority being of the form of deterministic systems of nonlinear differential equations [4, 5, 7, 29–31]). Here, too, a deterministic model will be developed and used to study the dynamics of the disease. A notable feature of the model to be developed is that it incorporates numerous pertinent aspects of the vaccination program and the current knowledge of the epidemiology of the COVID-19 pandemic, including the waning and boosting of both the vaccine-derived and natural immunity. The model will be parameterized using cumulative case data for the COVID-19 pandemic during the onset of the Omicron variant in the United States. The rest of the paper is organized as follows. The model is formulated, and fitted with observed COVID-19 case data, in Section 2. Basic qualitative features of the model are also given. Rigorous analysis of the model, with respect to the existence and asymptotic stability of its disease-free equilibrium, is carried out in Section 3. Expressions for vaccine-derived herd immunity thresholds are also derived in Section 3. Numerical simulations are carried out in Section 4.

## 2. Model Formulation

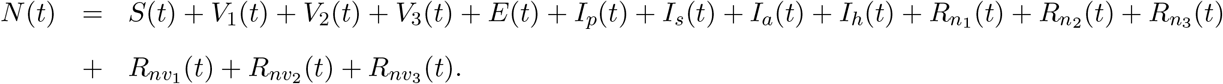

Numerous clinical studies show that the vaccine-derived immunity against SARS-CoV-2 begin to wane after nine months of the receipt of the full vaccine doses [27, 31, 32]. Consequently, in our model formulation, individuals in the *V*_1_ class (who enjoy high level of the protective efficacy of the vaccine) are those that are within nine months of receipt of full vaccine doses. Furthermore, individuals in the *V*_2_ class are those who have received the full doses between 9 months to a year ago (hence, the vaccine efficacy is moderate). Finally, individuals in the *V*_3_ class are assumed to have received the full vaccine doses at least 2 years ago (and the vaccine efficacy is very mild). This study allows for the waning and boosting of vaccine-derived and natural immunity (boosting of natural immunity is assumed to occur due to treatment or the use of other immune-boosting supplements [33, 34]).

The model is given by the following deterministic system of nonlinear differential equations, where a dot represents differentiation with respect to time *t* (a flow diagram of the model is depicted in Figure 1, and the state variables and parameters of the model are described in Tables 1 and 2, respectively):

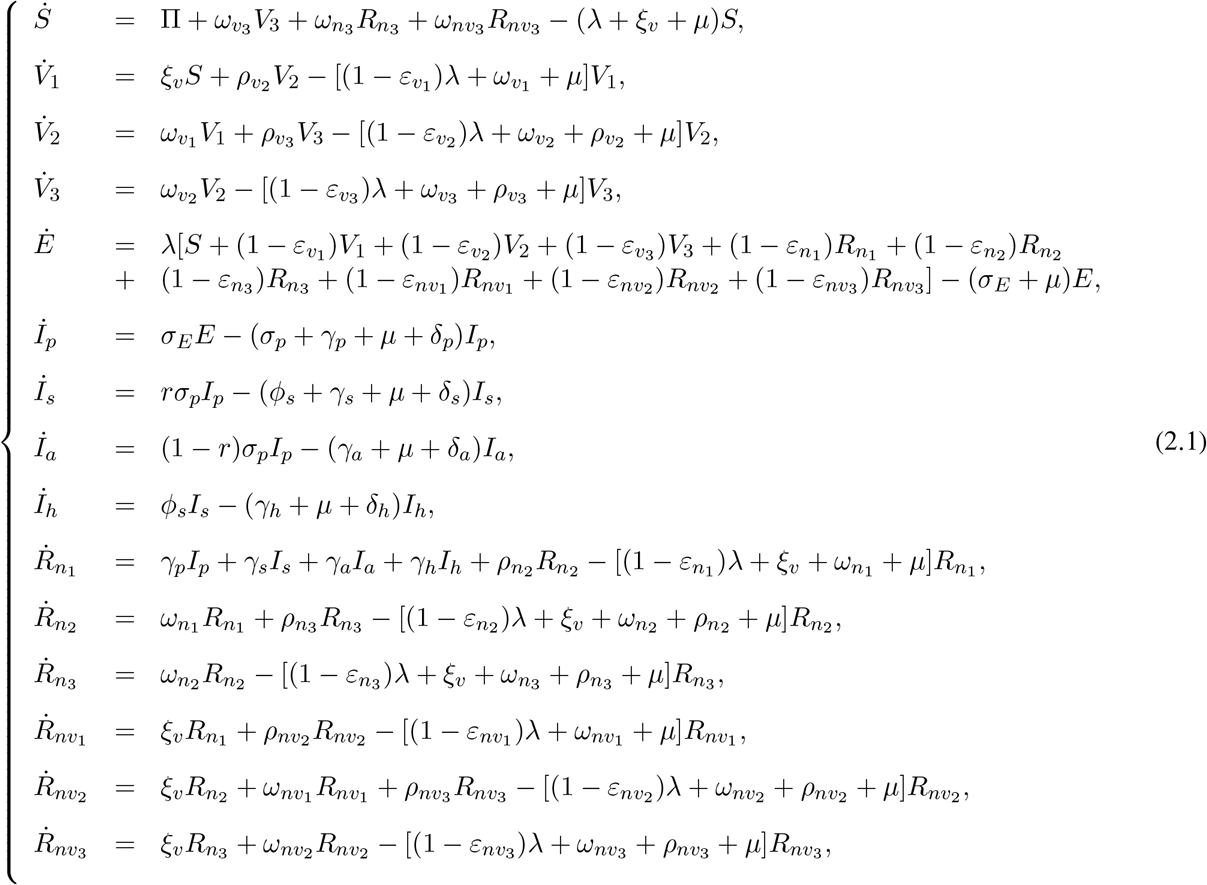

where,

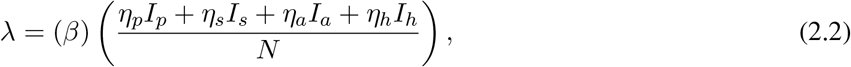

is the infection rate. In (2.2), *β* is the effective contact rate for individuals and *η*_*i*_ (with *i* = *p, s, a, h*) is the modification parameter for the heterogeneity in the infectiousness of infected individuals in the presymptomatic (*I*_*p*_), symptomatic (*I*_*s*_), asymptomatic (*I*_*a*_) and hospitalized (*I*_*h*_) class, respectively.

**Figure 1:**
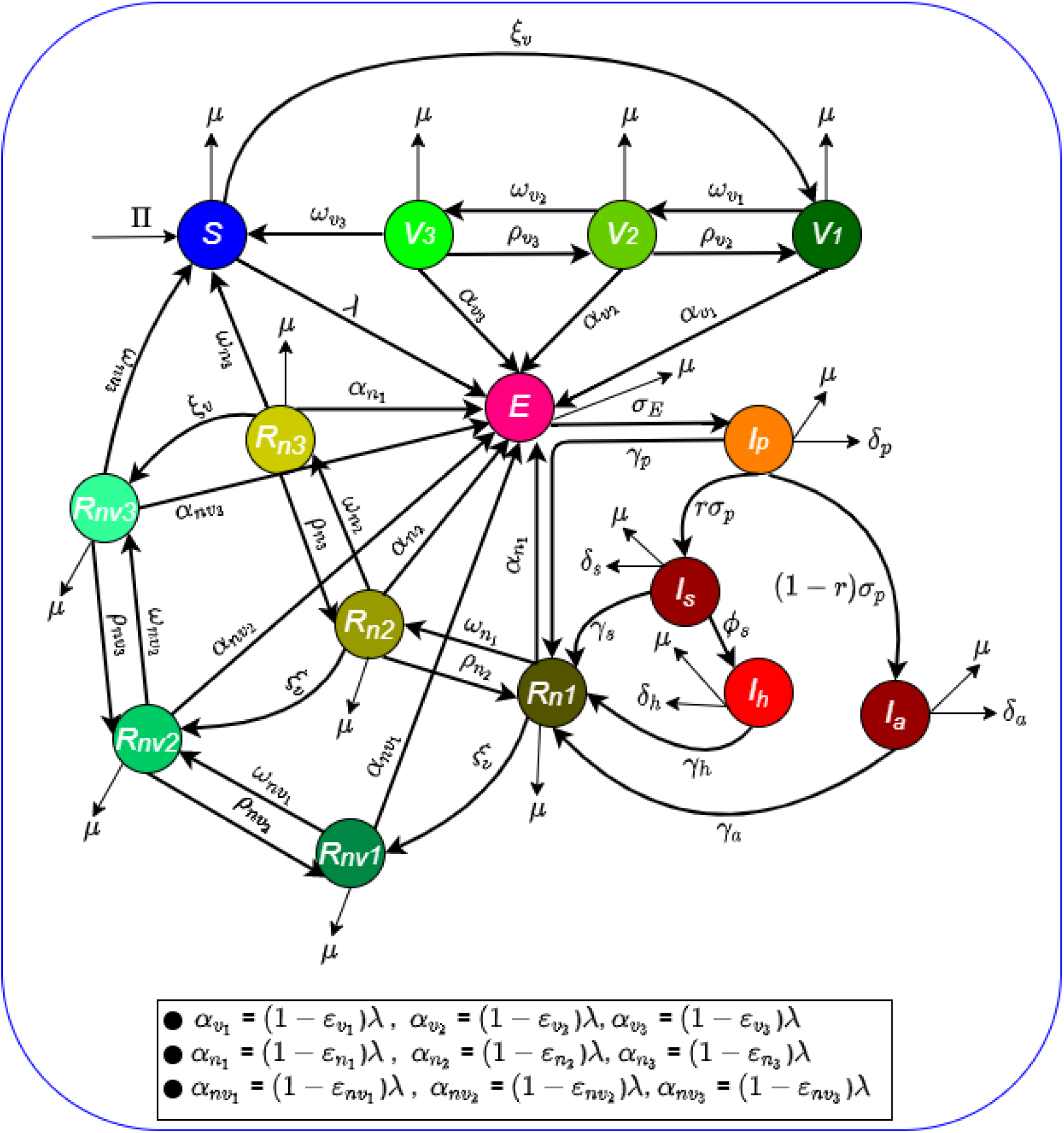
Flow diagram of the model (2.1).

**Table 1:** Description of the state variables of the model (2.1).

| State variable | Description |
| --- | --- |
| $S$ | Population of unvaccinated (wholly) susceptible individuals |
| $V_1$ | Population of vaccinated susceptible individuals with high vaccine-derived immunity |
| $V_2$ | Population of vaccinated susceptible individuals with moderate vaccine-derived immunity |
| $V_3$ | Population of vaccinated susceptible individuals with low vaccine-derived immunity |
| $E$ | Population of exposed (newly-infected individuals) |
| $I_p$ | Population of pre-symptomatic infectious individuals |
| $I_s$ | Population of infectious individuals with clinical symptoms of the disease |
| $I_a$ | Population of asymptotically-infectious individuals |
| $I_h$ | Population of hospitalized individuals |
| $R_{n1}$ | Population of recovered individuals with high natural immunity |
| $R_{n2}$ | Population of recovered individuals with moderate natural immunity |
| $R_{n3}$ | Population of recovered individuals with low natural immunity |
| $R_{nv1}$ | Population of recovered individuals with high natural and vaccine-derived immunity |
| $R_{nv2}$ | Population of recovered individuals with moderate natural and vaccine-derived immunity |
| $R_{nv3}$ | Population of recovered individuals with low natural and vaccine-derived immunity |

**Table 2:** Description of the parameters of the model (2.1).

| Parameter | Description |
| --- | --- |
| $\Pi$ | Recruitment rate |
| $\beta$ | Effective contact rate of individuals |
| $\eta_i (i = p, s, a, h)$ | Modification parameter for the infectiousness of the individuals in $I_p, I_s, I_a$, and $I_h$ classes |
| $\xi_v$ | Vaccination rate |
| $\mu$ | Natural death rate |
| $r$ | Proportion of individuals who show clinical symptoms of the disease |
| $\omega_{v_i} (i = 1, 2, 3)$ | Waning rate of vaccinated individuals in stage $V_i$ |
| $\omega_{n_i} (i = 1, 2, 3)$ | Waning rate of natural immunity in individuals in stage $R_{n_i}$ |
| $\omega_{nv_i} (i = 1, 2, 3)$ | Waning rate of natural plus vaccine-derived immunity in individuals in stage $R_{nv_i}$ |
| $\rho_{v2}(\rho_{v3})$ | Boosting rate of vaccine-derived immunity of the individuals in stage $V_2(V_3)$ |
| $\rho_{n2}(\rho_{n3})$ | Boosting rate of natural immunity of the individuals in stage $R_{n2}(R_{n3})$ |
| $\rho_{nv2}(\rho_{nv3})$ | Boosting rate of vaccine-derived and natural immunity of those in stage $R_{nv2}(R_{nv3})$ |
| $\varepsilon_{v_i} (i = 1, 2, 3)$ | Vaccine efficacy for vaccinated individuals in $V_1, V_2$ and $V_3$ classes, respectively |
| $\varepsilon_{n_i} (i = 1, 2, 3)$ | Efficacy of natural immunity to prevent infection of recovered individuals in $R_{n1}, R_{n2}$ and $R_{n3}$ |
| $\varepsilon_{nv_i} (i = 1, 2, 3)$ | Efficacy of natural and vaccine derived immunity to prevent infection of recovered individuals in $R_{nv1}, R_{nv2}$ and $R_{nv3}$ |
| $\sigma_E$ | Progression rate from exposed class to pre-symptomatic class |
| $\sigma_p$ | Progression rate from pre-symptomatic class to either symptomatic or asymptomatic class |
| $\gamma_i (i = p, s, a, h)$ | Recovery rate for individuals in the $I_p, I_s, I_a$ and $I_h$ classes, respectively |
| $\phi_s$ | Hospitalization rate of individuals with clinical symptoms of the disease |
| $\delta_i (i = p, s, a, h)$ | Disease-induced mortality rate for individuals in the $I_p, I_s, I_a$ and $I_h$ classes, respectively |

In the model (2.1), Π is the recruitment of individuals into the population, 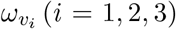 is the vaccine waning rate for vaccinated individuals in stage 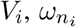 is the waning natural immunity for recovered individuals in stage 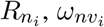 is the waning rate of both vaccine-derived and natural immunity for individuals in stage 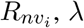 is the infection rate (defined in (2.2)), *ξ*_*v*_ is the *per capita* vaccination rate and *μ* is the natural death rate. Vaccinated individuals in *V*_2_ and *V*_3_ classes receive booster doses at the rate *ρ*_*vi*_ (*i* = 2, 3) and revert to the higher efficacy vaccination stage *V*_1_ and *V*_2_, respectively. Similarly, recovered individuals in the *R*_*n*2_ and *R*_*n*3_ classes receive immune booster at a rate *ρ*_*n*2_ and *ρ*_*n*3_, respectively (and revert, respectively, to stages *R*_*n*1_ and *R*_*n*2_). Individuals in *R*_*nv*2_ and *R*_*nv*3_ (that have both the vaccine-derived and natural immunity) receive a booster at a rate *ρ*_*nv*2_ and *ρ*_*nv*3_, respectively (and revert to *R*_*nv*1_ and *R*_*nv*2_, respectively).

The parameter *ε*_*vi*_ is the protective efficacy of the vaccine for vaccinated susceptible individuals in stage *V*_*i*_ (*i* = 1, …, 3), while 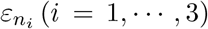 is the efficacy of natural immunity to prevent recovered individuals (in the *R*_*nvi*_ class) from acquiring future SARS-CoV-2 infection. of the recovered individuals. Exposed individuals progress to the pre-symptomatic stage at the rate *σ*_*E*_, and pre-symptomatic individuals progress to either become symptomatically-infectious, at a rate *rσ*_*p*_ (where 0 *≤ r ≤* 1 is the proportion of these individuals that show clinical symptoms), or become asymptomatically-infectious, at the rate (1 *− r*)*σ*_*p*_. Symptomatic individuals are hospitalized at a rate *ϕ*_*s*_, and infectious individuals in stage *I*_*i*_ (with *i* = *p, s, a, h*) recover at a rate *γ*_*i*_ (*i* = *p, s, a, h*). Finally, disease-induced mortality occur in the *I*_*p*_, *I*_*s*_, *I*_*a*_ and *I*_*h*_ classes at a rate *δ*_*i*_ (*i* = *p, s, a, h*).

Some of the main assumptions made in the formulation of the model (2.1) include:

a. Homogeneous mixing: we assumed a well-mixed population, such that every member of the community is equally likely to mix with (and acquire infection from) every other member of the community.
b. Vaccinated susceptible individuals (in the *V*_1_, *V*_2_ and *V*_3_ classes) are assumed to have received the full required doses (i.e., two doses for Pfizer or Moderna vaccine, one dose for the Johnson & Johnson vaccine), and that enough time has elapsed for the body to develop immunity.
c. The three SARS-CoV-2 vaccines that received FDA’s Emergency Use Authorization (Pfizer, Moderna and Johnson & Johnson) are imperfect [10, 13, 35]. That is, the vaccines offer partial protective immunity (with efficacy 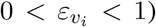, which wanes over time (at a rate 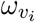), for *i* = 1, …, 3 [27, 32]. In other words, vaccinated individuals can experience breakthrough infection [36, 37].
d. We assumed gradual waning of both vaccine-derived and natural immunity over time, resulting, ultimately, in reverting to the wholly-susceptible class *S* [27]. Moreover, we assume that the average waiting times in the 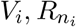 and 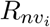 (with *i* = 1, …, 3) classes are gamma-distributed [38].
e. Vaccination is only offered to wholly-susceptible individuals or those who recovered naturally from COVID-19 infection but their natural immunity has waned completely or those recovered individuals who had acquired natural plus vaccine-derived immunity after recovering from COVID-19 infection but the immunity has completely waned over time. In other words, individuals who are currently infected are not vaccinated.
f. Immunity level can be increased or strengthened, by using immunity boosters [33, 34, 39, 40], for the individuals in the 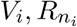 and 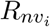 (*i* = 1, …, 3) classes.

### 2.1. Data Fitting and Parameter Estimation

In this section, we fit the model (2.1) by using the available data for cumulative COVID-19 case for the U.S. (for the period November 28, 2021–February 23, 2022). The model (2.1) has several parameters, some of which are known from the literature (as tabulated in Table (3)) and the remaining unknown parameters are obtained by fitting the model (2.1) with the cumulative case data obtained from the Johns Hopkins University COVID-19 repository [3]. The model was fitted using a standard nonlinear least squares approach, which involved using the inbuilt MATLAB minimization function “lsqcurvefit” to minimize the sum of the squared differences between each observed cumulative cases data points and the corresponding cumulative cases points obtained from the model (2.1) (i.e., *rσ*_*p*_*I*_*p*_). The unknown parameters which are estimated from the fitting are presented in Table 4.

**Table 3:** Baseline values of the fixed parameters of the model (2.1).

| Parameter | Value | Source | Parameter | Value | Source |
| --- | --- | --- | --- | --- | --- |
| $\sigma_E$ | $1/3 \text{ day}^{-1}$ | [52] | $\omega_{v_1}$ | $1/274 \text{ day}^{-1}$ | [53] |
| $\sigma_p$ | $1/2 \text{ day}^{-1}$ | [54] | $\omega_{v_2}$ | $1/365 \text{ day}^{-1}$ | [53] |
| $r$ | 0.099 (dimensionless) | [55] | $\omega_{v_3}$ | $1/365 \text{ day}^{-1}$ | [53] |
| $\phi_s$ | $1/5 \text{ day}^{-1}$ | [31] | $\omega_{n_1}$ | $1/274 \text{ day}^{-1}$ | [53] |
| $\gamma_s$ | $1/10 \text{ day}^{-1}$ | [27, 50] | $\omega_{n_2}$ | $1/365 \text{ day}^{-1}$ | [53] |
| $\gamma_a$ | $1/5 \text{ day}^{-1}$ | [27, 50] | $\omega_{n_3}$ | $1/365 \text{ day}^{-1}$ | [53] |
| $\gamma_h$ | $1/8 \text{ day}^{-1}$ | [27, 50] | $\omega_{nv_1}$ | $1/548 \text{ day}^{-1}$ | Assumed |
| $\eta_p$ | $5/4 \text{ day}^{-1}$ | [27] | $\omega_{nv_2}$ | $1/730 \text{ day}^{-1}$ | Assumed |
| $\eta_s$ | $1/2 \text{ day}^{-1}$ | [8] | $\omega_{nv_3}$ | $1/730 \text{ day}^{-1}$ | Assumed |
| $\eta_a$ | $3/2 \text{ day}^{-1}$ | [27] | $\rho_{v_2}$ | $1/14 \text{ day}^{-1}$ | [56] |
| $\eta_h$ | $3/20 \text{ day}^{-1}$ | [8] | $\rho_{v_3}$ | $1/14 \text{ day}^{-1}$ | [56] |
| $\xi_v$ | $1.9 \times 10^{-5} \text{ day}^{-1}$ | [57] | $\rho_{n_2}$ | $1/14 \text{ day}^{-1}$ | [56] |
| $\Pi$ | $11400 \text{ day}^{-1}$ | [8] | $\rho_{n_3}$ | $1/14 \text{ day}^{-1}$ | [56] |
| $\mu$ | $3.4 \times 10^{-5} \text{ day}^{-1}$ | [8] | $\delta_h$ | $5.0 \times 10^{-5} \text{ day}^{-1}$ | [8] |

**Table 4:** Baseline values of fitted (estimated) parameters of the model (2.1), obtained by fitting the model with the observed cumulative daily COVID-19 data for the United States for the period November 28th, 2021 to February 23rd, 2022.

| Parameter | Estimated Value | Parameter | Estimated Value |
| --- | --- | --- | --- |
| $\beta$ | $0.2120 \text{ day}^{-1}$ | $\rho_{nv_2}$ | $0.1996 \text{ day}^{-1}$ |
| $\rho_{nv_3}$ | $0.6398 \text{ day}^{-1}$ | $\delta_s$ | $4.9804 \times 10^{-5} \text{ day}^{-1}$ |

The data fitting is done by splitting the available COVID-19 cumulative case data for the United States for the period from November 28, 2021 (when Omicron first emerged) to March 23, 2022 into two segments. The first segment of the data, from November 28, 2021 to February 23, 2022 (i.e., the region to the left of the dashed vertical cyan line), was used to fit the model (2.1) and to estimate the unknown parameters. The results obtained, depicted in Figure 2 (a), show a very good fit for the model output (blue curve) and the observed daily case data (red dots). The data for the second segment of the data, for the period from February 24th, 2022 to March 23, 2022, was used for the cross validation of the fitted data. This also shows a very good fit for the model output (green curve) and for the remaining data points of the observed data (red dots) of Figure 2 (a). This segment of the Figure 2 (a) clearly shows that the model (2.1) cross validates the observed cumulative case data for the period from February 23, 2022 to March 23, 2022 perfectly (solid green curve). Furthermore, we show, in this figure, the prediction of the model for the cumulative COVID-19 cases for approximately a five-week period after March 24, 2022 (i.e., the region to the right of the dashed vertical black line), as illustrated by the solid magenta curve in Figure 2 (a). The model was then simulated (using the fixed and fitted parameter baseline values in Tables 5, 3 and 4), and compared with the observed new daily cases data. The results obtained, depicted in Figure 2 (b), show a very good fit for the daily new COVID-19 cases in the United States.

**Figure 2:**
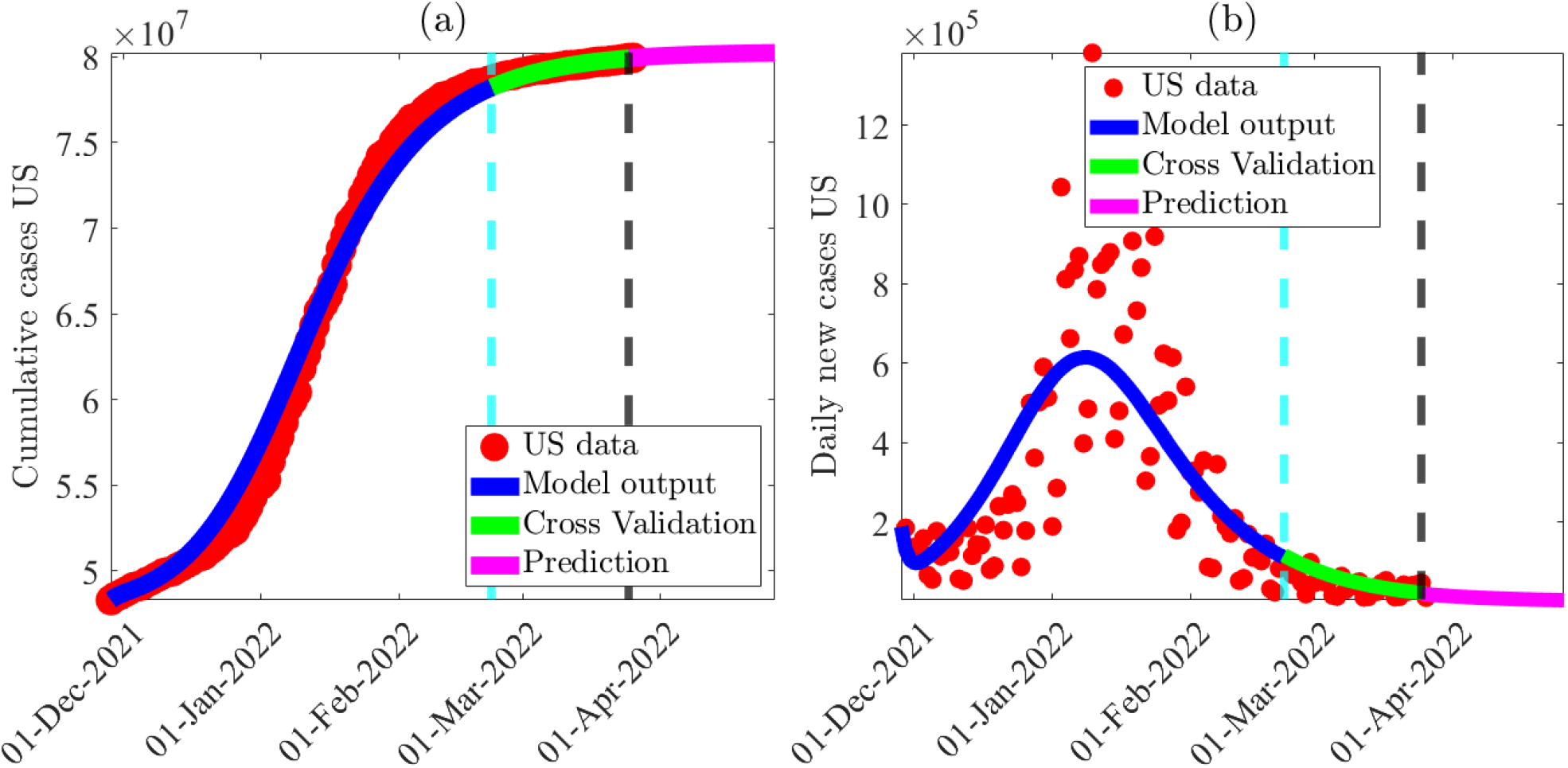
(a) Time series illustration of the least squares fit of the model (2.1), showing the model’s output for the cumulative daily cases in the United States (blue curve) compared to the observed cumulative confirmed cases for the United States (red dots) from November 28, 2021 to February 23, 2022 (segment to the left of the dashed vertical cyan line). (b) Simulation result of (2.1), showing observed daily confirmed COVID-19 cases for the United States as a function of time, using the fixed and estimated baseline parameter values given in Tables 5, 3 and 4, respectively. The segment from February 24, 2022 to April 30, 2022 (i.e., solid green and magenta curves or the entire segment to the right of the dashed cyan vertical line) illustrates the performance of the model (2.1) in predicting the cumulative and daily cases in the United States.

**Table 5:** Assumed baseline levels of the parameters for the efficacy of the vaccine-derived and natural immunity.

| Vaccine Efficacy of $V_n$ class | Vaccine Efficacy of $R_n$ class | Vaccine Efficacy of $R_{nv}$ class |
| --- | --- | --- |
| $\varepsilon_{v_1} = 0.85$ | $\varepsilon_{n_1} = 0.85$ | $\varepsilon_{nv_1} = 0.95$ |
| $\varepsilon_{v_2} = 0.50$ | $\varepsilon_{n_2} = 0.50$ | $\varepsilon_{nv_2} = 0.50$ |
| $\varepsilon_{v_3} = 0.20$ | $\varepsilon_{n_3} = 0.20$ | $\varepsilon_{nv_3} = 0.20$ |

### 2.2. Basic Qualitative Analysis

Before carrying out the asymptotic analysis and numerical simulations of the model (2.1), it is instructive to explore its basic qualitative features with respect to its well-posedness (i.e., with respect to the non-negativity, boundedness and invariance of its solutions). First of all, since the model (2.1) monitors the temporal dynamics of human populations, all its parameters are non-negative. It is convenient to define the following biologically-feasible region for the model: (2.1):

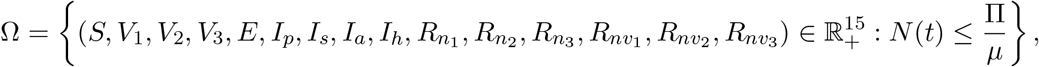

where *N* (*t*) is the total population. For the model (2.1) to be mathematically- and biologically-meaningful, it is necessary that the solutions of the model (2.1) remain non-negative for all non-negative initial conditions. That is, solutions that start in Ω remain in Ω for all time *t >* 0 (i.e., Ω is positively-invariant with respect to the model (2.1)). Furthermore, let

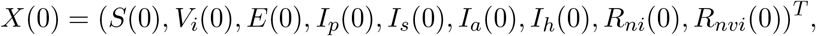

with *i* = 1, 2, 3, be the vector of initial solutions of the model (2.1). We claim the following result.

#### Theorem 2.1.

*Consider the model* (2.1) *with non-negative initial data X*(0). *The region* Ω *is positively-invariant and bounded with respect to the model* (2.1).

*Proof*. Adding all the equations of the model (2.1) gives

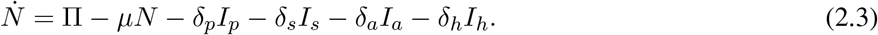

By the non-negativity of parameters for model (2.1), it follows from (2.3) that

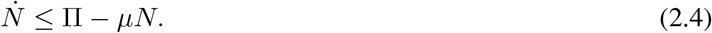

Hence, if 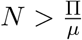, then 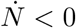. Thus, it follows, by applying a standard comparison theorem [41] on (2.4), that:

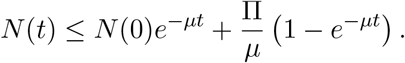

Hence, if 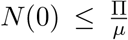, then 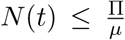. Thus, the solutions of the model (2.1) are bounded. Therefore, every solution of the model (2.1) with initial conditions in Ω remains in Ω for all time *t*. In other words, the region Ω is positively-invariant and attracts all initial solutions of the model (2.1).

The consequence of Theorem 2.1 is that it is sufficient to consider the dynamics of the flow generated by (2.1) in Ω, since the model (2.1) is epidemiologically and mathematically well-posed [42] there.

## 3. Asymptotic Stability Analysis of Disease-free Equilibria

The disease-free equilibrium of the model (2.1) is given by:

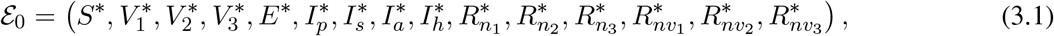

where,

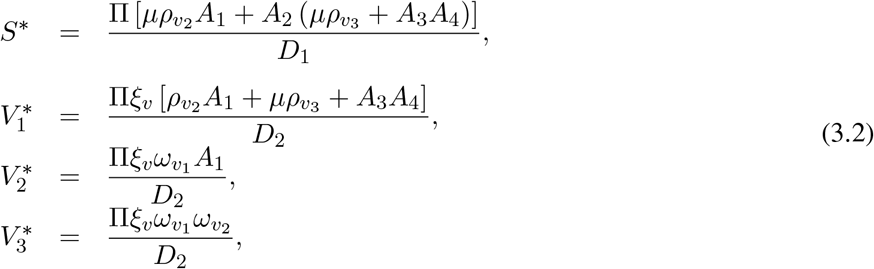

with,

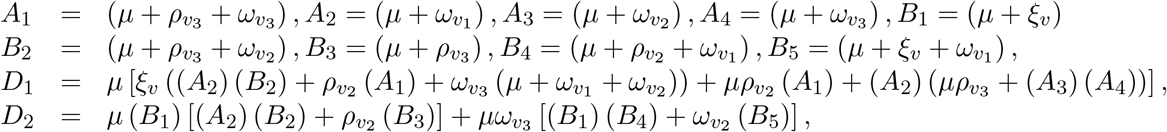

and all other components (for the infected and recovered compartments of the model) take the value zero.

The asymptotic stability property of the DFE (*ℰ*_0_) can be explored using the *next generation operator method* [43, 44]. Using the notation in [43], it follows that the associated non-negative matrix of new infection terms (*F*) and the M-matrix of the linear transition terms (*V*) are given, respectively, by:

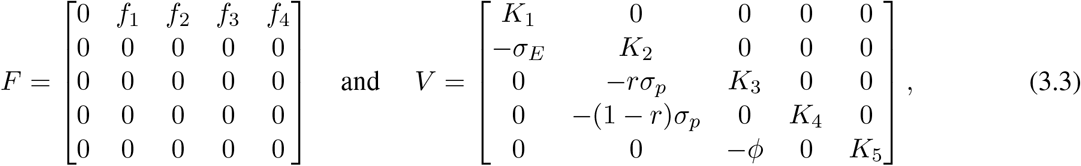

Where 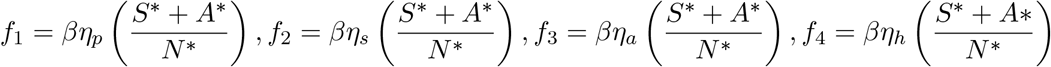, with

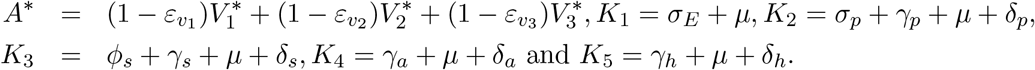

It is convenient to define the quantity (where *ρ* is the spectral radius):

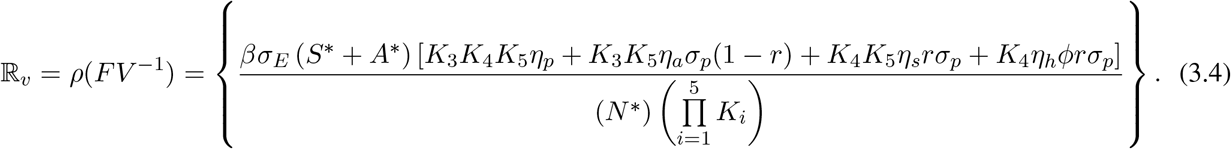

The result below follows from Theorem 2 of [43].

### Theorem 3.1.

*The disease-free equilibrium* (*E*_0_) *of the model* (2.1) *is locally-asymptotically stable (LAS) if* ℝ_*v*_ *<* 1, *and unstable if* ℝ_*v*_ *>* 1.

The threshold quantity ℝ_*v*_ is the *vaccination reproduction number* of the model (2.1), which measures the average number of new COVID-19 cases generated by a single infectious individual introduced into a population where a certain proportion is vaccinated. The epidemiological interpretation of Theorem (3.1) is that a small influx of COVID-19 cases will not generate a large outbreak in the community if the vaccination reproduction number (ℝ_*v*_) is brought to, and maintained at a, value less than unity.

In the absence of vaccination and other public health interventions, the vaccination reproduction number (ℝ_*v*_) reduces to the basic reproduction number (denoted by ℝ_0_). That is,

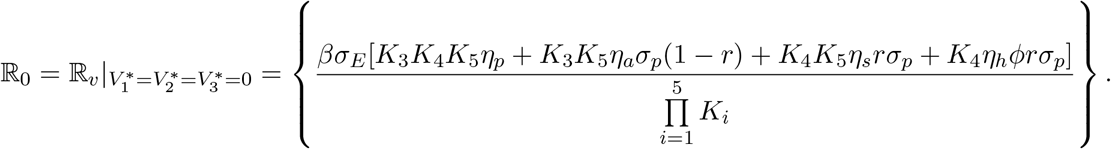

### 3.1. Global asymptotic stability of DFE: Special Case

Consider the special case of the model (2.1) with perfect vaccine protective efficacy against primary infection and re-infection and no waning of vaccine-derived and natural immunity (i.e., we consider the model (2.1) with 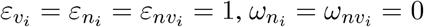, with *i* = 1, 2, 3). It is convenient to let:

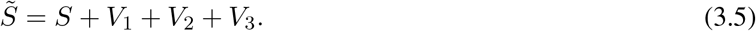

Substituting 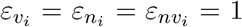 and 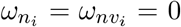 (with *i* = 1, 2, 3) into the model (2.1), it follows that the equation for the rate of change of the new compartment 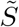 (defined in (3.5)) is given by (where the infection rate, *λ*, is as defined in (2.2)):

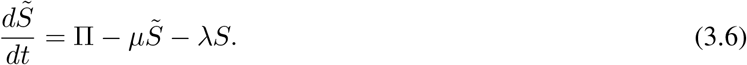

It can be shown that the special case of the model is positively-invariant and bounded in the region (as shown in Section 2.1)

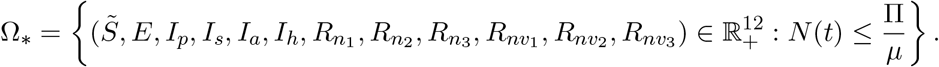

Furthermore, the disease-free equilibrium of the special case of the model is given by:

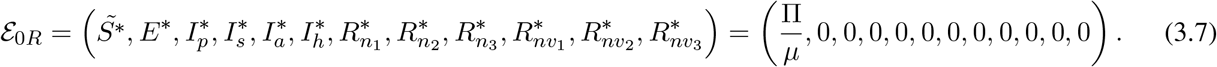

For the aforementioned special case of the model, it can be seen that the associated next generation matrix of new infection terms, denoted by 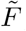, is given by (note that, for this special case, the next generation matrix of linear transition terms, *V*, remains the same, as given in (3.3). Further, *N*^*∗*^ = Π*/μ*):

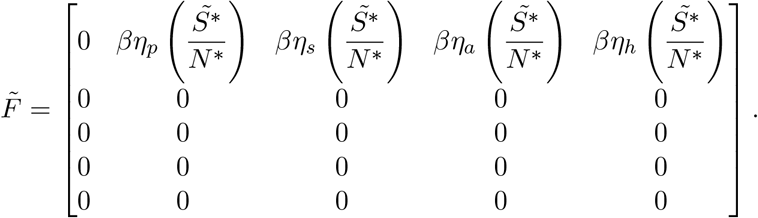

Thus,

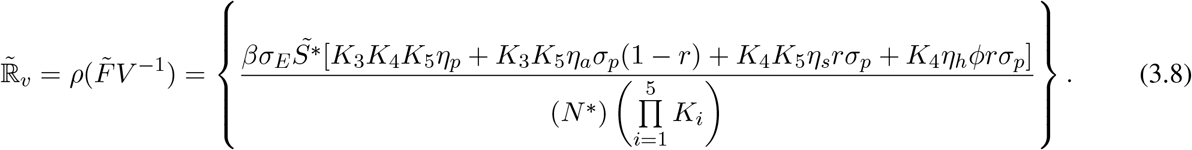

We claim the following result:

#### Theorem 3.2.

*Consider the special case of the model* (2.1) *with* 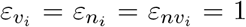 *and* 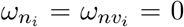 *(for i* = 1, 2, 3*). The disease-free equilibrium of the special case of the model* (*ℰ* _0*R*_) *is globally-asymptotically stable in* Ω_*∗*_ *whenever* 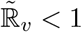.

*Proof*. Consider the model (2.1) with 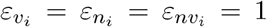 and 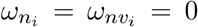 (*i* = 1, 2, 3). Further, let 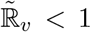. The proof is based on using a comparison theorem [45]. It can be shown, first of all, that the region Ω_*∗*_ is positively-invariant and attracts all solutions of the aforementioned special case of the model (2.1) [46] (as shown in Section 2.1). The equations for the infected compartments of the special case of the model (2.1) can be re-written in terms of the next generation matrices (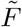 and *V*) as below:

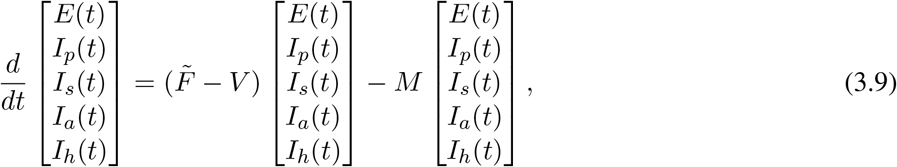

where,

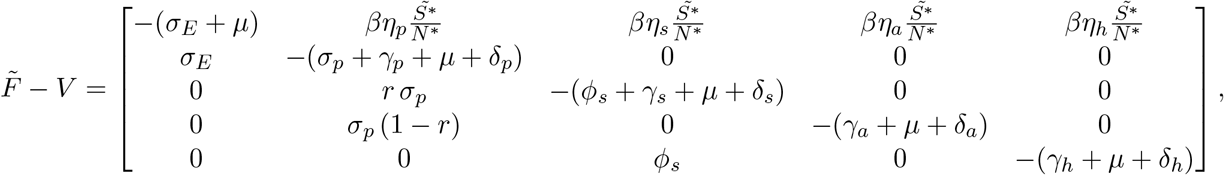

and,

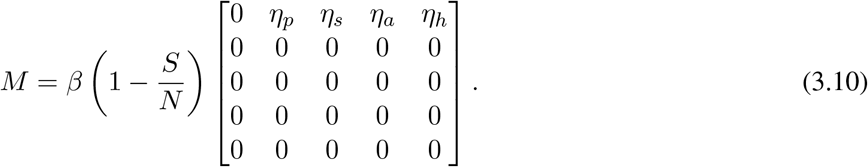

Since *S*(*t*) *≤ N* (*t*) for all *t >* 0 in Ω_*∗*_, it follows that the matrix *M*, defined in (3.10), is non-negative. Hence, the equation (3.9) can be re-written in terms of the following inequality:

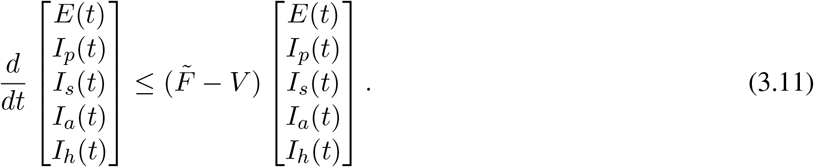

It should be recalled from the local asymptotic stable result for the DFE (given in Theorem 3.1) that all eigenvalues of the next generation matrix 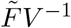 are negative if 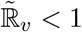 (i.e., 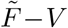 is a stable matrix). Thus, it can be concluded that the linearized differential inequality system (3.11) is stable whenever 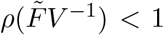. Hence, it follows that (for the linear system of ordinary differential equations (3.11)):

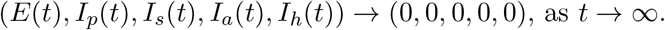

Substituting *E*(*t*) = *I*_*p*_(*t*) = *I*_*s*_(*t*) = *I*_*a*_(*t*) = *I*_*h*_(*t*) = 0 into the differential equations for the rate of change of the 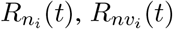 and 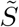 (with *i* = 1, 2, 3) compartments of the model (2.1) shows that (where 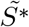 is defined in (3.7)):

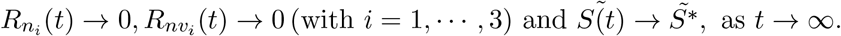

Thus, the DFE (*ℰ*_0*R*_) of the special case of the model (2.1), with 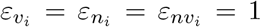 and 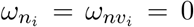 (*i* = 1, 2, 3), is globally-asymptotically stable in Ω_*∗*_ whenever 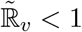.

Epidemiologically-speaking, Theorem 3.2 shows that, for the special case of the model (2.1) with 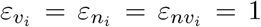 and 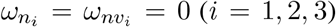, the disease can be eliminated from the community if the threshold quantity, 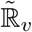, can be brought to (and maintained at) a value less than unity.

The global asymptotic stability of the disease-free equilibrium of the model (2.1) can also be established for another special case, as described below. Consider the special case of the model (2.1) in the absence of disease-induced mortality (i.e. *δ*_*p*_ = *δ*_*s*_ = *δ*_*a*_ = *δ*_*h*_ = 0) and no reinfection (i.e., 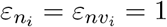 with *i* = 1, 2, 3). Setting *δ*_*p*_ = *δ*_*s*_ = *δ*_*a*_ = *δ*_*h*_ = 0 in the model (2.1), and adding all the equations of the model shows that 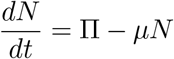, from which it follows that 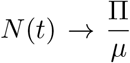 as *t* → ∞. From now on, we replace *N* (*t*) with its limiting value, *N*^*∗*^ = Π*/μ* (i.e., the standard incidence formulation for the infection rate is now replaced by a mass action incidence). Furthermore, it is convenient to define the following feasible region for the special case of the model (where *S*^*∗*^ and 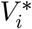, with *i* = 1, 2, 3, are as defined in Section 3):

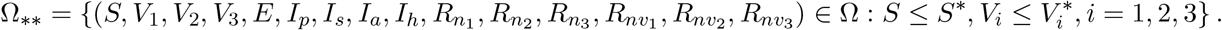

It can be shown that the region Ω_*∗∗*_ is positively-invariant with respect to the aforementioned special case of the model [46]. Further, for this special case of the model, it can be shown that the associated next generation matrices are given, respectively, by:

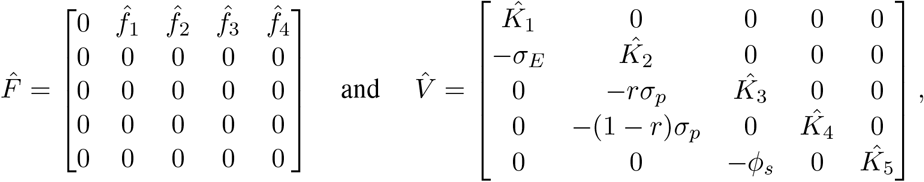

where (with *S*^*∗*^ and *A*^*∗*^ as defined in Section 3),

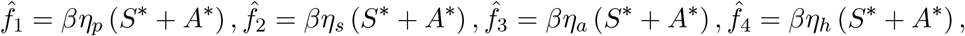

and,

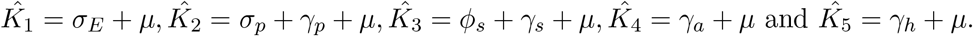

It is convenient to define the following threshold quantity:

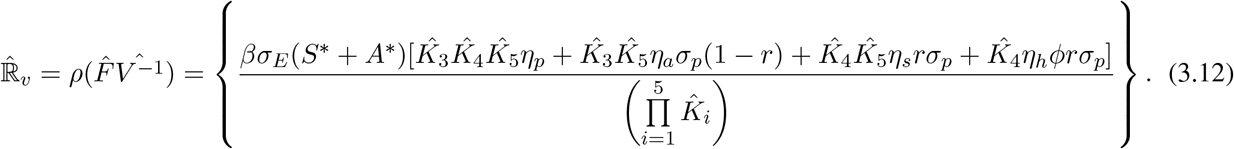

We claim the following result:

#### Theorem 3.3.

*Consider the special case of the model* (2.1) *in the absence of disease-induced mortality (i.e., δ*_*p*_ = *δ*_*s*_ = *δ*_*a*_ = *δ*_*h*_ = 0) and no reinfection of recovered individuals (*i.e*., 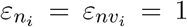 *with i* = 1, 2, 3*). The disease-free equilibrium of the special case of the model* (ℰ_0_) *is globally-asymptotically stable in* Ω_*∗∗*_ *whenever* 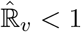.

The proof of Theorem 3.3, based on using a comparison theorem, is given in Appendix A.

### 3.2. Derivation of Vaccine-Induced Herd Immunity Threshold

Herd immunity, which is a measure of the minimum percentage of the number of susceptible individuals that need to be protected against the infection in order to eliminate community transmission of an infectious disease, can be attained through two main ways, namely natural immunity route (following natural recovery from infection with the disease) or by vaccination (which is widely considered to be the safest and the fastest way) [47, 48]. For vaccine-preventable diseases, such as COVID-19, it is not practically possible to vaccinate every susceptible individual in the community due to various reasons, such as individuals with certain underlying medical conditions, infants, individuals who are pregnant, breastfeeding women or those who are unwilling to be vaccinated for COVID-19 due to some other reasons [29]. Therefore, it is crucial to determine the minimum proportion of the susceptible population that need to be vaccinated in order to protect those that cannot be vaccinated (so that vaccine-induced herd immunity is achieved in the population). Since we have three vaccination classes (*V*_1_, *V*_2_ and *V*_3_), accounting for the three levels of vaccine-derived immunity (high, moderate and low), we will compute vaccine-derived herd immunity thresholds for the United States with respect to each of the vaccination classes. Specifically, we let

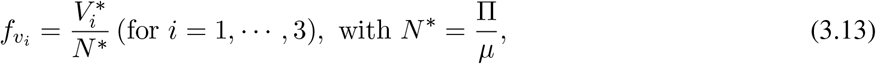

represent the proportion of susceptible members of the population that have been fully-vaccinated (using any of the three approved vaccines) at the disease-free equilibrium (*ℰ*_0_). Using the definition (3.13) in (3.4) gives:

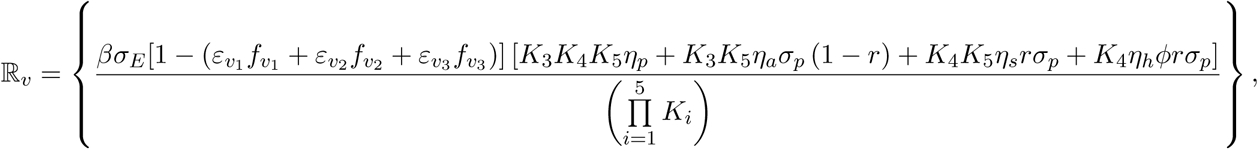

which can be expressed in terms of the basic reproduction number (ℝ_0_) as:

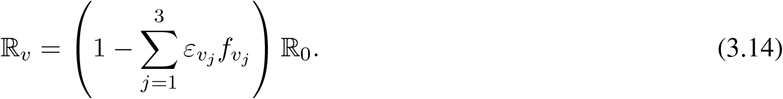

Setting ℝ_*v*_ = 1 (i.e., the bifurcation point) in Equation (3.14), and simplifying, gives:

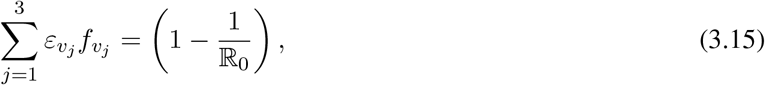

from which we can solve for the fraction fully-vaccinated (those in *V*_1_ class), or received booster doses (i.e., those in *V*_2_ and *V*_3_ classes), at steady-state for each vaccinated class (denoted by 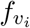 with *i* = 1, …, 3), in terms of the basic reproduction number, giving:

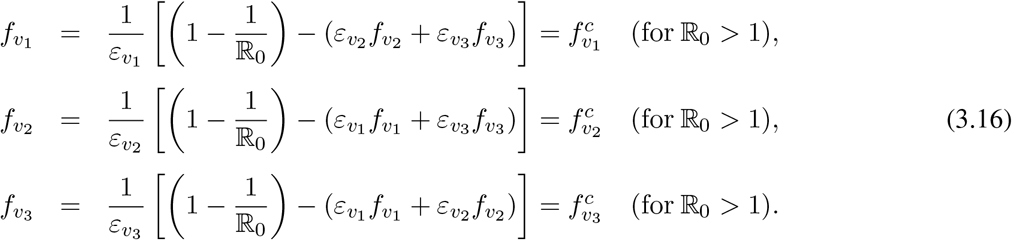

It follows from Equation (3.16) that 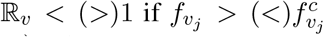 (with *i* = 1, …, 3). Furthermore, ℝ_*v*_ = 1 whenever 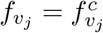 (with *i* = 1, …, 3). This result is summarized below:

#### Theorem 3.4.

*Vaccine-induced herd immunity (i*.*e*., *COVID-19 elimination) can be achieved in the United States, using any of the approved anti-COVID vaccine, if vaccination of susceptible individuals and boosting of vaccine-derived immunity resulted in* 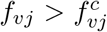 *(i.e., if* ℝ_*v*_ < 1*) for each corresponding j, for all j* = 1, 2, 3. *If* 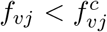 *(i.e*., *if* ℝ_*v*_ *>* 1*), then the vaccination program will fail to eliminate the COVID-19 pandemic*.

Epidemiologically-speaking, Theorem 3.4 implies that the use of any of the approved COVID-19 vaccines can lead to the elimination of the pandemic in the United States if the proportion of susceptible individuals fully-vaccinated and with high level of vaccine-derived immunity (i.e., those in *V*_1_ class) and boosted (i.e., those in *V*_2_ and *V*_3_ classes) at steady-state reached or exceeded the aforementioned critical threshold values. In other words, the SARS-CoV-2 pandemic will be eliminated in the United States if 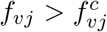 for each corresponding *j*, for all *j* = 1, 2, 3. On the contrary, the Vaccination program will fail to eliminate the pandemic if the proportion vaccinated (and boosted) at the disease-free equilibrium falls below the aforementioned critical herd immunity thresholds.

It should be mentioned that since the Pfizer and Moderna vaccines offer protective efficacy of about 95% and 94%, respectively [49], and the Johnson & Johnson vaccine offers a protective efficacy of about 67% [50], we set the average vaccine protective efficacy for individuals in the *V*_1_ class to be

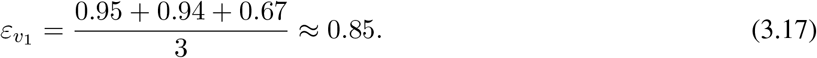

Table 5 summarizes the assumed baseline efficacy levels for vaccine-derived and natural immunity to be used in our numerical simulations. Using the baseline values of the fixed and fitted parameters in Tables 3-4, together with the baseline vaccine-derived and natural immunity protective efficacy levels in Table 5, it follows from Equation (3.16) that the critical vaccine-derived herd immunity threshold for each of the vaccinated compartment is given, respectively, by 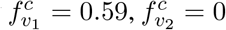 and 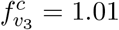. In other words, based on the parameterization of the model (2.1) with the recent case data for Omicron BA.1 variant in the United States, population-level herd immunity can be achieved in the United States if the following conditions hold:

a. at least 59% of the wholly-susceptible individuals are fully vaccinated (i.e., 59% of individuals in the *S* class are fully-vaccinated and moved to the *V*_1_ class);
b. 42% of vaccinated individuals with moderate vaccine-derived immunity (i.e., those in *V*_2_ class) are boosted;
c. almost all of the vaccinated individuals whose level of vaccine-derived immunity is low (i.e., those in *V*_3_) are boosted.

Hence, vaccine-derived herd immunity will be achieved *via* the aforementioned vaccination-boosting strategy that entails having at least 59% of the wholly-susceptible population to be fully-vaccinated followed by the boosting of an average of (42%+101%)/2=71.5% of the fully-vaccinated individuals with moderate and low vaccine-derived immunity.

Figure 3 depicts contour plots of the vaccination reproduction number (ℝ_*v*_), as a function of vaccination efficacy (*ε*_*vi*_) and coverage of fully-vaccinated or boosted individuals at steady-state (*f*_*vi*_), for *i* = 1, 2, 3. It follows from these plots that, for the overall vaccine-protective efficacy set at 85% (as stated above), at least 59% of the wholly-susceptible population need to be vaccinated at steady-state to bring the vaccination reproduction number (ℝ_*v*_) below one (Figure 3 (a)). For the case when the vaccine protective efficacy has waned to 50% (i.e., fully-vaccinated individuals now have moderate vaccine-derived immunity; here, *ε*_*v*2_ = 0.5, as given in Table 5), up to 42% of individuals in the *V*_2_ class need to be boosted to bring the vaccination reproduction number to a value below one (Figure 3 (b)). Finally, when vaccine-derived immunity has waned to the low level of 20%, the contour plot in Figure 3 (c) shows that all of the fully-vaccinated individuals with low vaccine-derived immunity (i.e., individuals in the *V*_3_ class) need to be boosted to bring the reproduction number to a value less than one. In summary, the results depicted in Figure 3 show that population-level herd immunity can be achieved in the United States *via* the implementation a vaccination program (based on using any of the three approved vaccines) that emphasizes the full vaccination of a sizable proportion of the susceptible pool (at least 59%) followed by the administration of booster doses to individuals in whom their vaccine-derived immunity has waned to moderate (at least 42%) or low (at least 100%) levels. Overall, our study shows that, for the case where the protective immunity offered by the vaccine for fully-vaccinated individuals in the *V*_1_ class is 85%, vaccine-derived herd immunity can be achieved in the United States if at least 59% of the susceptible population is fully-vaccinated (with any of the three approved vaccines) followed by the boosting of at least 71.5% of the fully-vaccinated individuals whose vaccine-derived immunity has waned to moderate or low level.

**Figure 3:**
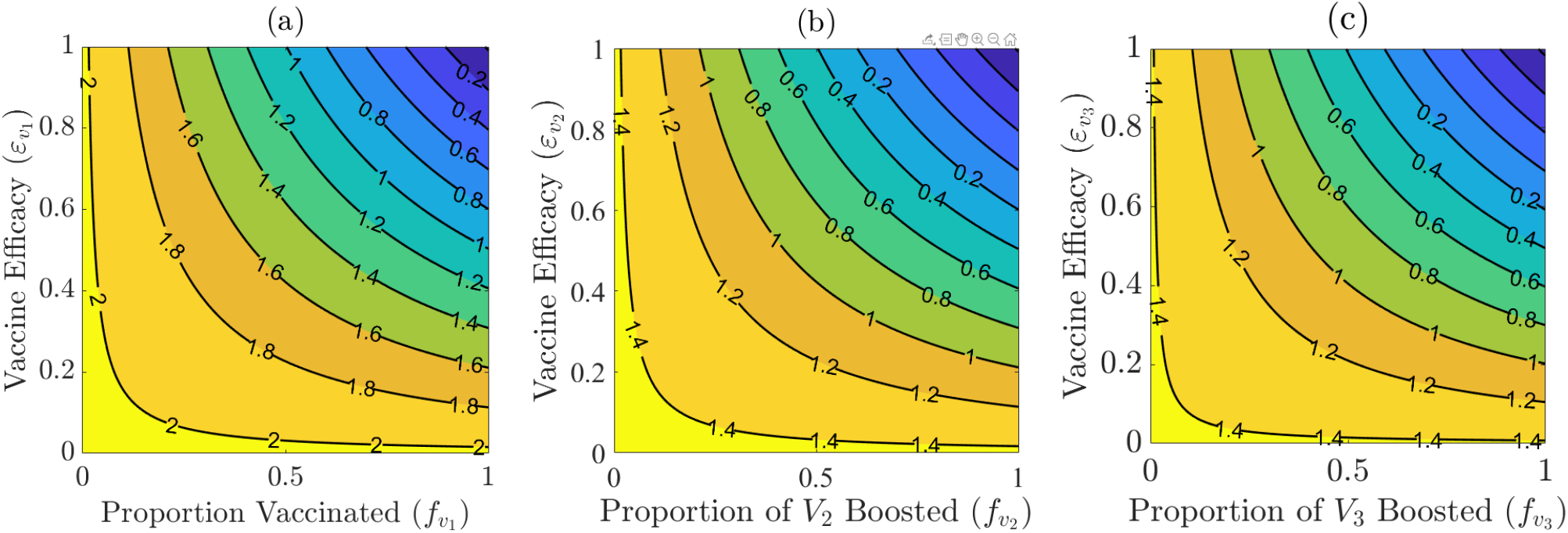
Contour plots of the vaccine reproduction number (ℝ_*v*_) of the model (2.1), as a function of vaccine coverage or boosting at steady-state 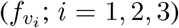 and vaccine efficacy 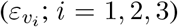, for the United States. (a) Vaccination of wholly-susceptible individuals (*S*(*t*); *f*_*v*1_ is proportion of wholly-susceptible individuals who are fully-vaccinated at steady-state). (b) Boosting of vaccinated individuals with moderate vaccine-derived immunity (*V*_2_(*t*); *f*_*v*2_ is the proportion of vaccinated individuals in the *V*_2_ class who are boosted). (c) Boosting of vaccinated individuals with low vaccine-derived immunity (*V*_3_(*t*); *f*_*v*3_ is the proportion of vaccinated individuals in the *V*_3_ class who are boosted). Parameter values used in these simulations are as given by their respective baseline values in Tables 3-5.

It should be mentioned that for the case when the high level of the vaccine-induced efficacy for individuals in the *V*_1_ class is decreased to 55%, for instance (while the vaccine protective efficacy for individuals in the *V*_2_ and *V*_3_ classes remain at the baseline level), our simulations showed that at least 91% of the wholly-susceptible population need to be fully-vaccinated, followed by the (marginal) boosting (about 4%) of the vaccinated individuals whose vaccine-derived immunity has waned to moderate or low level. Thus, this study shows that lower protective efficacy of the vaccine (for fully-vaccinated individuals) incurs higher requirement for the vaccination coverage of the susceptible population (followed by a correspondingly low level of boosting for the vaccinated individuals whose vaccine protective efficacy has waned to moderate or low level) to achieve herd immunity. Vaccinating 91% of the wholly-susceptible population is, of course, not realistically feasible in large populations, such as the United States. Hence, it is imperative that highly efficacious vaccines are developed and used, and combined with boosting (at moderate to high levels) of vaccinated individuals whose immunity has waned to moderate or low level. In other words, using vaccines with higher protective efficacy (e.g., vaccines with 85% protective efficacy, as computed in Equation (3.17)) incurs lower, and realistically attainable, requirement for the vaccination coverage (about 59%) and attainable (moderate to high) boosting level (about 71.5%) for the fully-vaccinated individuals whose vaccine-derived immunity has waned to moderate or low level. As of May 20, 2022, data from the CDC shows that about 66.5% of the U.S. population is fully-vaccinated, and about 46.4% of the population of fully-vaccinated individuals is boosted [51]. Thus, this study shows that, even for the scenario where the three vaccines offer such high protective efficacy, a sizable proportion of the fully-vaccinated individuals need to be boosted in order to achieve vaccine-derived herd immunity in the United States (using the aforementioned combined vaccination-boosting strategy).

## 4. Numerical Simulations

The model (2.1) will now be simulated to assess the population-level impact of waning and boosting of vaccine-derived and natural immunity on the dynamics of the Omicron variant in the United States. Unless otherwise stated, the simulations will be carried out using the baseline values of the parameters tabulated in Tables 3-5.

### 4.1. Assessing the impact of waning of vaccine-derived immunity: with and without boosting

To assess the impact of waning of vaccine-derived immunity for this scenario, we simulate the model (2.1) using the following three (arbitrarily-chosen) levels of the parameters related to the waning of vaccine-derived immunity in the population:

i. Low (i.e., slow) level of waning of vaccine-derived immunity: here, we consider vaccine-derived immunity to wane within 48 months (i.e., we set 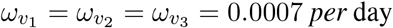) but parameters related to natural immunity and combined natural and vaccine-derived immunity are maintained at baseline level.
ii. Baseline level of waning of vaccine-derived immunity: in this case, waning of vaccine-derived immunity is set to occur within 9 months (so that, 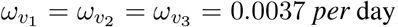) but parameters related to natural immunity and combined natural and vaccine-derived immunity are maintained at baseline level.
iii. High (i.e., fast) level of waning of vaccine-derived immunity: in this scenario, it is assumed that vaccine-derived immunity wanes within 3 months (i.e., 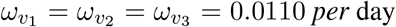), but parameters related to natural immunity and combined natural and vaccine-derived immunity are maintained at baseline level.

For these simulations, all other parameters of the model (including those that involve the waning of natural immunity, as stated above) are maintained at their baseline values (given in 3-5). Furthermore, these simulations are carried in the absence and presence of boosting of vaccine-derived immunity (recall that boosting of vaccine-derived immunity, maintained at baseline level, is achieved *via* the administration of the required doses of any of the approved SARS-CoV-2 booster vaccines used in the United States).

The simulation results obtained, depicted in Figure 4. First of all, these simulations also depict the fitting of the model’s output for the daily new and cumulative cases with the observed data (used in Section 2.1) for the baseline scenario (as shown by the blue curves and the red dots in Figure 4). Furthermore, these simulations show that, in the absence of boosting of vaccine-derived immunity, waning of vaccine-derived immunity generally induces only a marginal impact on the average number of new daily COVID-19 cases in the United States, for each of the three waning levels considered in our simulations, in comparison to the baseline scenario. For example, under the fast waning scenario for vaccine-derived immunity (i.e., vaccine-derived immunity wanes within three months, but natural immunity is maintained at its baseline level) and no boosting of vaccine-derived immunity is implemented, the simulations show a marginal (about 2%) increase in the peak level of the daily new cases, in comparison to the peak baseline level (this is evident by comparing the blue and magenta curves in Figure 4 (*a*), and the zoomed-in version of the segments of the curves near the peaks shown in Figure 4 (*b*)). For the slow waning scenario (i.e., if the vaccine-derived immunity wanes within 48 months, but natural immunity is maintained at baseline level), the increase in daily new cases at the peak (in comparison to the baseline) reduces to about 1.5% (compare the blue and green curves in Figures 4 (*a*) and (*b*)).

**Figure 4:**
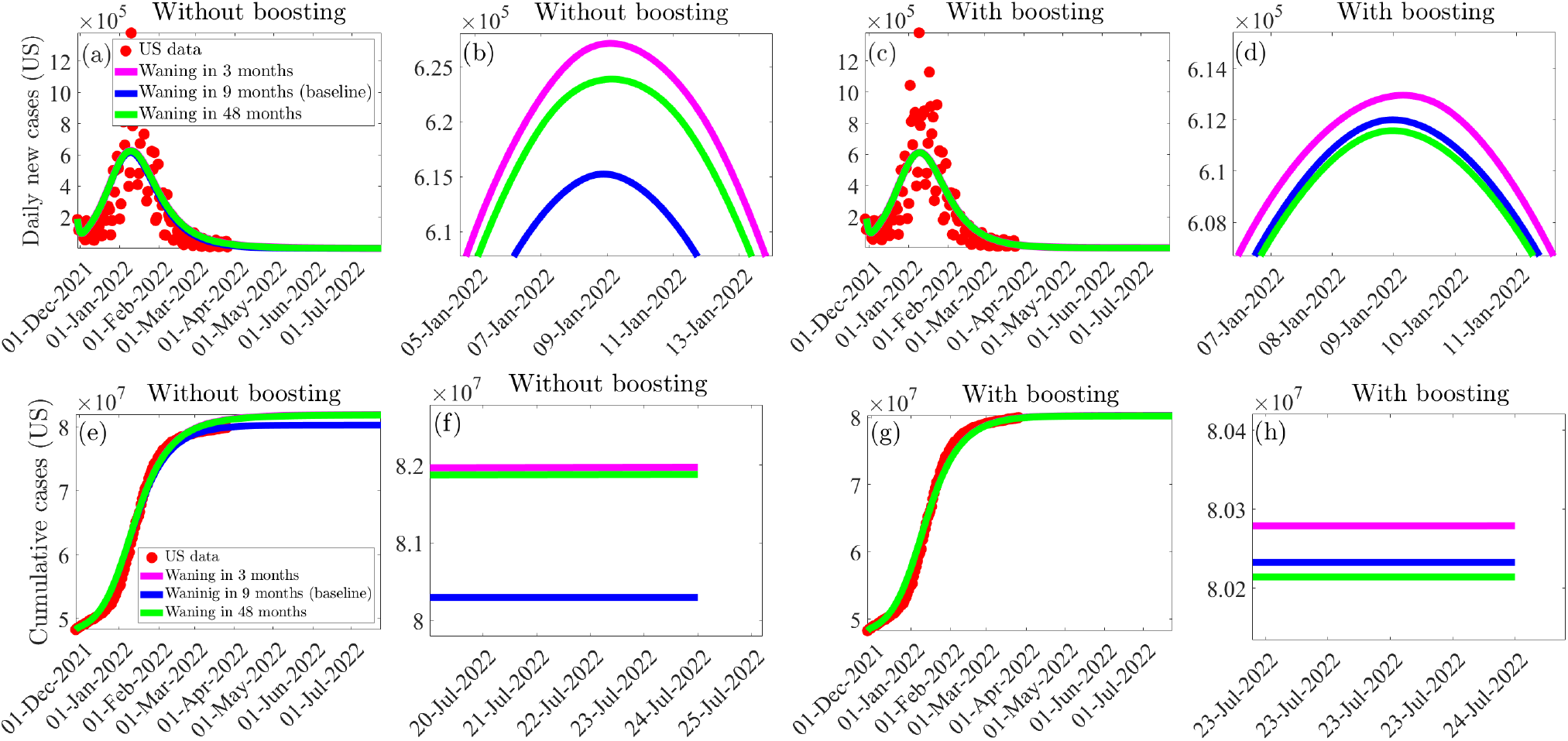
Simulations of the model (2.1) to assess the population-level impact of waning of vaccine-derived immunity in the absence and presence of boosting of vaccine-derived immunity (maintained at baseline level). (*a*) *−* (*d*): average number of new daily cases at the peak in the absence ((*a*) and (*b*)) and presence ((*c*) and (*d*)) of boosting of vaccine-derived immunity. (*e*) *−* (*h*): cumulative number of new cases in the absence ((*e*) and (*f*)) and presence ((*g*) and (*h*)) of boosting of vaccine-derived immunity (maintained at baseline level). Three levels of waning of vaccine-derived immunity were considered: vaccine-derived immunity wanes in three months (magenta curves), nine months (blue curves) and forty eight months (green curves). Zoomed-in versions of the portions of the curves near the peaks depicted in Figures (*a*) and (*c*) are shown in Figures (*b*) and (*d*), respectively. Similarly, zoomed-in versions of the portions of the curves near the peaks in Figures (*e*) and (*g*) are shown in Figures (*f*) and (*h*), respectively. The values of the other parameters of the model used in these simulations are as given in Tables 3-5.

In the presence of boosting of vaccine-derived immunity (at baseline level), our simulations show a significant reduction in the increase in the average number of daily new cases at the peak recorded under the above waning scenarios without boosting of vaccine-derived immunity. For instance, for the case where vaccine-derived immunity wanes within three months (but natural immunity is maintained at baseline level), boosting of vaccine-derived immunity at the baseline level significantly reduces the increase in daily new cases at the peak (by about 90%), in comparison to the corresponding case without boosting of vaccine-derived immunity (compare the blue and magenta curves in Figure 4 (*c*), and the corresponding zoomed-in portions of the curves near the peaks shown in Figure 4 (*d*)). Furthermore, under the slow waning scenario, boosting of vaccine-derived immunity at baseline level further increases the reduction in the peak daily new cases (compare the green and blue curves in Figures 4(*c*) and (*d*)). Similar dynamics are observed (and illustrated) with respect to the cumulative number of new cases, for the three waning scenarios considered in these simulations without (Figures 4 (*e*) and (*f*)) and with (Figures 4 (*g*) and (*h*)) boosting of vaccine-derived immunity.

We further simulated the model to assess the impact of waning and boosting of vaccine-derived immunity (for the case where natural immunity is maintained at baseline) for the following two scenarios:

#### Scenario (a): Fast waning and slow boosting

Here, we assume that the waning of vaccine-derived immunity range between 3 to 6 months and the duration of boosting of vaccine-derived immunity range from 20 days to 180 days.

#### Scenario (b): Fast waning and boosting near the baseline level

Under this scenario, vaccine-derived immunity wanes within the same 3 to 6 months period (as in Scenario (a)), but boosting of vaccine-derived immunity is accelerated to be implemented within 10 to 20 days (i.e. near the baseline level of 14 days).

The results obtained are depicted in the form of heat maps for the vaccination reproduction number (ℝ_*v*_) of the model (2.1), as a function of the rates of waning (*ω*_*v*_) and boosting (*ρ*_*v*_) of vaccine-derived immunity in Figure 5. This figure shows that, for the fast waning and slow boosting scenario (i.e., Scenario (a)), the values of the vaccination reproduction number lie in the range ℝ_*v*_ *∈* [0.82, 1.21] (with a mean of ℝ_*v*_ *≈* 1.015), suggesting that the disease will persist in the population (this is in line with the theoretical result given in Theorem 3.1). In other words, this result shows that faster waning and slower boosting, in comparison to waning and boosting at baseline levels, increases the prospect for disease persistence in the population. For Scenario (b), our simulations (Figure 5 (*b*)) show a marked decrease in the range of the reproduction number, with ℝ *∈* [0.78, 0.89] (with a mean of ℝ_*v*_ = 0.835), suggesting possible elimination of the pandemic (in line with Theorems 3.1-3.3). Thus, boosting of vaccine-derived (near the baseline rate) enhances the prospect for pandemic elimination.

**Figure 5:**
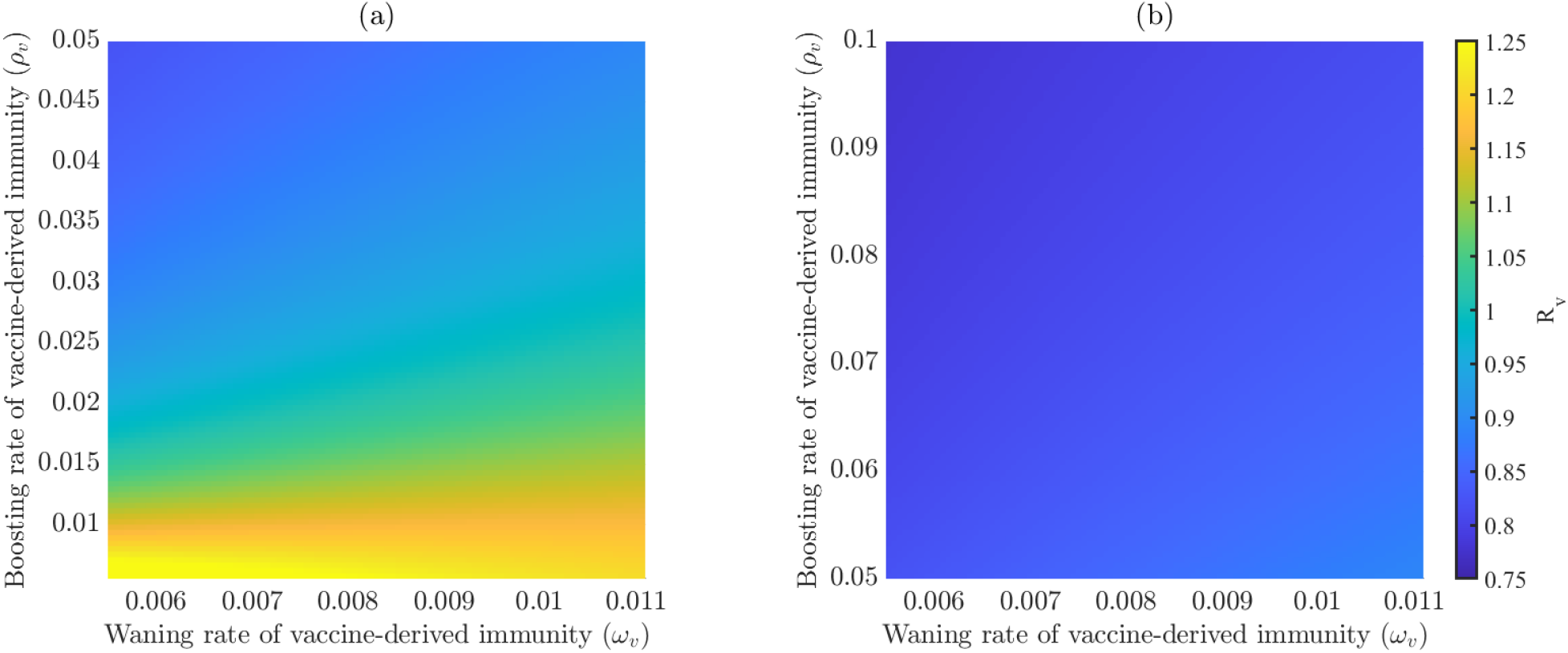
Effect of waning and boosting of vaccine-derived immunity. Heat maps of the vaccination reproduction number (ℝ_*v*_), as a function of the rates of waning (*ω*_*v*_) and boosting (*ρ*_*v*_) of vaccine-derived immunity. (*a*) Waning of vaccine-derived immunity range between 3 to 6 months, and duration of boosting of vaccine-derived immunity range from 20 days to 180 days (slow boosting). (*b*) Waning of vaccine-derived immunity range from 3 to 6 months, while duration of boosting of vaccine-derived immunity range from 10 to 20 days (fast boosting).

In summary, while the simulations in this section show that waning of vaccine-derived immunity generally induces only a marginal impact in the average number of new cases at the peak of the COVID-19 pandemic, boosting of vaccine-derived immunity (maintained at its baseline level) resulted in a dramatic reduction in the average number of new cases at the peak, in comparison to the case where boosting is not implemented. Furthermore, delay in boosting of vaccine-derived immunity, in comparison to the baseline level of boosting, could alter the trajectory disease from possible elimination (as measured by the vaccine reproduction number, ℝ_*v*_, taking a value less than one) to persistence of the disease (as measured by the reproduction number being greater than one).

### 4.2. Assessing the effect of waning of natural immunity: with and without boosting

Natural immunity can be boosted *via* treatment or the use of other immune-boosting supplements [33, 34]). To assess the impact of waning of natural immunity, we simulated the model (2.1) using the following (arbitrarily-chosen) waning levels:

i. Low (i.e., slow) level of waning of natural immunity: here, too, we consider natural immunity to wane within 48 months (i.e., we set 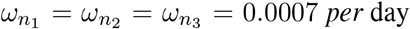), but vaccine-derived immunity and combined natural and vaccine-derived immunity are kept at baseline.
ii. Baseline waning of natural immunity: in this case, waning of natural immunity is set to occur within 9 months (so that, 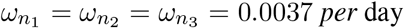), but vaccine-derived immunity and combined natural and vaccine-derived immunity are kept at baseline.
iii. High (i.e., fast) level of waning of natural immunity: here, too, natural immunity is assumed to wane within 3 months (i.e., 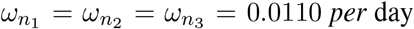), but vaccine-derived immunity and combined natural and vaccine-derived immunity are kept at baseline.

For the simulations in this section, we set all other parameters (including those related to the waning of vaccine-derived immunity and combined waning of natural and vaccine-derived) to their baseline values (given in Table 3-5). The simulation results obtained, depicted in Figure 6, also showed that waning of the natural immunity general only induces a marginal increase in the average number of new daily COVID-19 cases in the United States, in comparison to the baseline scenario (where the waning of natural immunity is assumed to occur within 9 months). In particular, if natural immunity wanes within three months and no boosting of natural immunity is implemented, the average number of new daily cases at the peak increases by about 4.2%, in comparison to the baseline scenario (compare the blue and magenta curves in Figure 6 (*a*), and the zoomed-in portions of the curves near the peaks, depicted in Figure 6 (*b*)). An additional marginal increase in the average number of new daily cases at the peak is recorded under the slow waning scenario for the natural immunity, in comparison to the baseline scenario (compare the blue and green curves in Figures 6 (*a*) and (*b*)).

**Figure 6:**
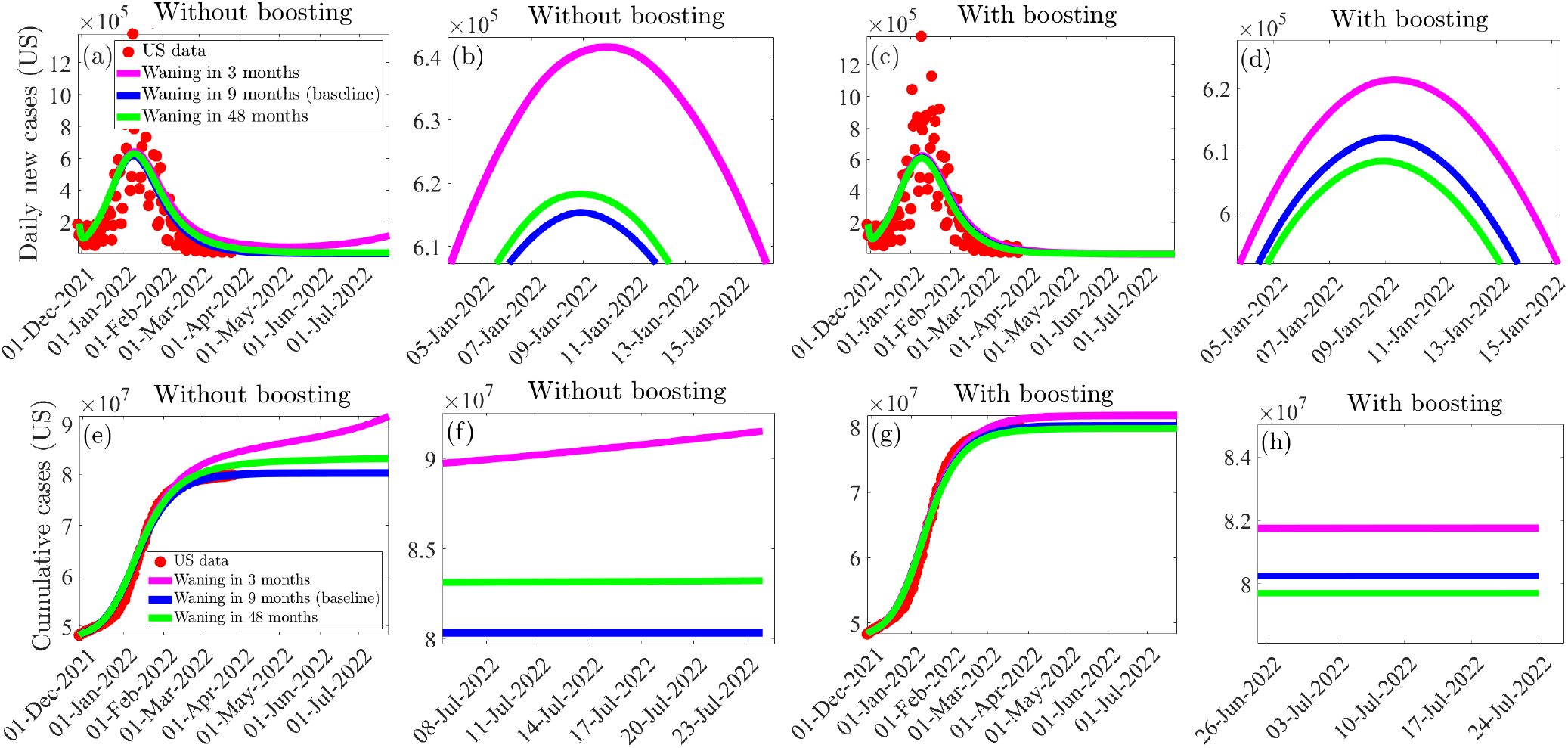
Simulations of the model (2.1) to assess the population-level impact of waning of natural immunity for the case with and without boosting of natural immunity (at the baseline level). (*a*) *−* (*d*): average number of new daily cases at the peak in the absence ((*a*) and (*b*)) presence ((*c*) and (*d*)) of boosting of natural immunity. (*e*) *−* (*h*): cumulative number of new cases in the absence ((*e*) and (*f*)) and presence ((*g*) and (*h*)) of boosting of natural immunity (maintained at baseline level). Three levels of waning of natural immunity were considered: natural immunity wanes in three months (magenta curves), nine months (blue curves) and forty eight months (green curves). Zoomed-in versions of the portions of the curves near the peaks depicted in Figures (*a*) and (*c*) are shown in Figures (*b*) and (*d*), respectively. Similarly, zoomed-in versions of the portions of the curves near the peaks in Figures (*e*) and (*g*) are shown in Figures (*f*) and (*h*), respectively. The values of the other parameters of the model used in these simulations are as given in Tables 3-5.

However, if natural immunity is boosted (at baseline level), our simulations show a marked reduction in the increase in the average daily new cases recorded at the peak, in comparison to the corresponding scenario without boosting of the natural immunity. Specifically, when natural immunity wanes within three months and boosting of natural immunity is implemented (maintained at its baseline level), the increase in the average number of new daily cases at the peak (in comparison to the baseline) reduces to about 1.6% (compare the blue and magenta curves in Figure 6 (*c*), and the zoomed-in portions near the peak depicted in Figure 6 (*d*)). This represents an approximately 62.2% reduction in the average daily new cases at the peak, in comparison to the corresponding scenario where natural immunity is not boosted. It should be mentioned that boosting of vaccine-derived immunity (at baseline) plays a more significant role in reducing the average number of new daily cases, in comparison to the corresponding boosting of natural immunity (this can be seen by comparing the corresponding peaks in Figures 4 and 6). In particular, while boosting of vaccine-derived immunity (at baseline) will lead to about 90% reduction in the number of new daily cases at the peak, boosting of natural immunity (at baseline) will lead to about 62% reduction in the number of new daily cases at the peak). Further significant reductions in the average number of new daily cases are recorded if the natural immunity wanes at a slower rate (compare the blue and green curves in Figures 6 (*a*) and (*c*) or (*b*) and (*d*), without and with boosting of natural immunity). We illustrated similar dynamics with respect to the cumulative number of new cases without (Figures 6 (*e*) and (*f*)) and with (Figures 6 (*g*) and (*h*)) boosting of natural immunity.

In summary, like in the case of waning of vaccine-derived immunity discussed in Section 4.1, the simulations in this section show that while the waning natural immunity only causes a marginal increase in the average number of new cases at the peak, boosting natural immunity (at baseline) resulted in a significant reduction in the average number of new cases recorded at the peak, in comparison to the scenario where a strategy for boosting of natural immunity is not implemented in the community.

## Discussion and Conclusions

The COVID-19 pandemic, caused by SARS-CoV-2, has made a significant impact on public health and the economy of almost every nation on earth since its emergence in December of 2019. The United States became the epicenter of the pandemic since late May, 2020 (recording the highest numbers of cumulative cases, hospitalizations and deaths). As of mid July, 2022, the virus had caused over 88.6 million and 1 million deaths in the United States [2, 3]. The rapid development, deployment, and administration of several safe and highly effective vaccines contributed significantly in curbing the spread of the virus worldwide. Three of these vaccines (the Pfizer-BioNTech, Moderna and Johnson & Johnson vaccines) have been approved by the FDA for use in the United States. The effectiveness of these vaccines in combating COVID-19 has been negatively affected by the emergence of various variants of SARS-CoV-2 (e.g., the Delta and Omicron variants). In particular, the Omicron (B.1.1.529) variant was declared a variant of concern by the World Health Organization in late November, 2021 [58], due to its exceptionally high transmissibility. Although all the available vaccines were developed for the original SARS-CoV-2 virus strain, they have been able to offer some level of cross-protection against other variants of concern. Furthermore, multiple studies have shown that the efficacy of vaccine-derived immunity wanes over time [27, 32, 59]. In order to overcome the waning effect of vaccine-derived immunity, booster vaccines were recommended by the CDC in November 2021 [39, 60].

In this study, we developed a mathematical model to assess the population-level impact of the waning and boosting of vaccine-derived and natural immunity against the Omicron BA.1 variant of SARS-CoV-2 in the United States. The model was parameterized by fitting it to the observed cumulative COVID-19 case data for the United States for the period from November, 28, 2021 to February 23, 2022 [3]. We used the remaining segment of the available data (i.e., the segment from February 24, 2022 to March 23, 2022) to cross validate the model. This cross validation, together with simulations involving the new daily COVID-19 cases, showed a good match to the observed data.

The model was rigorously analyzed to gain qualitative insight into the dynamics and burden of the diseases. The analysis showed that the disease-free equilibrium (DFE) of the model is locally-asymptotically stable whenever the vaccination reproduction number (denoted by ℝ_*v*_ *<* 1) is below one. Using the baseline values of the fixed and estimated parameters of the model, we computed the numerical value of R_*v*_ during the period of the emergence and circulation of the Omicron variant (starting from late November of 2021). The computed value was ℝ_*v*_ = 0.81 (suggesting that Omicron was on a downward trajectory towards elimination in the United States). The numerical value of the basic reproduction number of the model (which is ℝ_*v*_ *<* 1 computed in the absence of any control measure implemented) was ℝ_0_ = 2.051. We showed that the disease-free equilibrium of the model is globally-asymptotically for two special cases ((a) when the vaccines offer 100% protection against acquisition of infection and no reinfection and waning of immunity occurs and (b) disease-induced mortality is negligible and reinfection does not occur) when the associated vaccination reproduction number is less than one. The epidemiological implication of this global asymptotic stability result is that the SARS-CoV-2 pandemic can be eliminated if the the associated vaccination reproduction number can be brought to (and maintained at) a value less than one (in other words, having the value of this reproduction threshold less than one is necessary and sufficient for the elimination of the pandemic in the United States).

Explicit expression for the vaccine-induced herd immunity threshold was derived, and we showed, using current data for COVID-19 cases in the United States, that, for the case where the three vaccines offer 85% protective efficacy against the Omicron variant, vaccine-derived herd immunity will be achieved in the United States *via* a combined vaccination-boosting strategy that entails fully-vaccinating 59% of the wholly-susceptible population combined with the boosting of at least 71.5% of the population of the fully-vaccinated individuals whose vaccine-derived immunity has waned to moderate or low level. On the other hand, if the protective efficacy offered by the three vaccines is reduced to a lower level, such as 55% (as against 85% above), at least 91% of the wholly-susceptible population need to be vaccinated to achieve herd immunity. This very high level of vaccination coverage is not realistically attainable, especially in a large populations such as that of the United States. Data related to COVID-19 from the CDC show that, as of May 20, 2022, about 66.5% of the U.S. population was fully-vaccinated and 46.4% of this population received a booster [51]. Thus, our study suggests that, for the scenario that the three vaccines offer the reasonably high protective efficacy of 85% against acquisition of infection, herd immunity can realistically achieved by fully-vaccinating a moderate proportion (about 59%) of the wholly-susceptible and boosting about 71.5% of this cohort in whom the vaccine-derived immunity has waned to moderate or low level.

We conducted extensive numerical simulations to assess the impact of waning and boosting of vaccine-derived and natural immunity for each three arbitrarily selected waning scenarios (slow, baseline, and fast). Our study showed, based on these simulations, that in the absence of boosting of vaccine-derived and natural immunity, waning of vaccine-derived and natural immunity only causes a marginal increase in the average number of daily cases (at the peak) and the number of cumulative COVID-19 cases, in comparison to the baseline scenario. In other words, we showed that waning of either vaccine-derived or natural immunity (or both) has only marginal impact, for each of the three waning scenarios we considered, on the dynamics of the SARS-CoV-2 pandemic (as measured in terms of increases in the average number of daily new cases recorded at the peak, in comparison to the case where baseline values of all the parameters of the model are used).

We also showed that if fully-vaccinated individuals with moderate or low level of vaccine-derived immunity are boosted (at baseline level), the effect of waning of immunity is a lot less pronounced, in comparison to the baseline scenario (in other words, dramatic reductions in the increase in the average number of daily new cases at the peak recorded (under the three waning scenarios) are achieved if both immunity types are boosted at baseline level, in comparison to the corresponding scenarios where the immunity wanes but no boosting is implemented. We further showed that boosting of vaccine-derived immunity is more beneficial (in reducing average number of new cases) than boosting of natural immunity. Specifically, for the fast waning scenario, boosting of vaccine-derived immunity (at baseline level) resulted in an approximate 90% reduction in the average number of new daily cases at the peak, while boosting of natural immunity resulted in about *≈* 62% reduction in the number of new daily cases at the peak (in comparison to the corresponding scenarios without boosting). Furthermore, this study shows that boosting of vaccine-derived immunity (implemented near the baseline level) increased the prospects of altering the trajectory of the COVID-19 pandemic from persistence to possible elimination (even for the fast waning scenario of the vaccine derived-immunity) of the pandemic in the United States. Thus, the implementation of vaccination-boosting strategy greatly enhances the prospects of eliminating the COVID-19 pandemic in the United States.

In addition to the standard assumptions on which the model is built, some of the limitations of this study include the fact that we did not explicitly account for the impact of other control interventions (notably, the use of face masks, voluntary testing and detection of SARS-CoV-2 cases, isolation of confirmed cases, etc.), which also play important roles in the battle against the COVID-19 pandemic. Furthermore, this study assumes that the population is well-mixed and does not explicitly account for a number of heterogeneities, including age and risk structure, which may be relevant to gain insight into the dynamics of the disease. Furthermore, the current study did not account for the effects of other SARS-CoV-2 variants, including the BA.2 Omicron variant (which is more contiguous than the original BA.1 Omicron variant) [19, 26]. We fitted our deterministic model to cumulative case data instead of raw (new daily case) data, which might lead to narrower confidence intervals and/or provide a misleading measure of uncertainty. Hence, the results of the study should be interpreted with some caution. Nonetheless, our study shows, overall, that the prospect for the effective control and mitigation (and, consequently, elimination) of the COVID-19 pandemic in the United States is very promising using a combined vaccination-boosting strategy, provide the vaccinate and boosting coverages are moderately high enough.

## Data Availability

All data produced in the present study are available upon reasonable request to the authors

## Acknowledgements

ABG acknowledges the support, in part, of the Simons Foundation (Award #585022) and the National Science Foundation (Grant Number: DMS-2052363). CNN acknowledges the support of the Simons Foundation (Award #627346) and the National Science Foundation (Grant Number: DMS #2151870). SS acknowledges the support of the Fulbright Foreign Student Program.

## Appendix A. Proof of Theorem 3.3

*Proof*. Consider the special case of the model (2.1) with *δ*_*p*_ = *δ*_*s*_ = *δ*_*a*_ = *δ*_*h*_ = 0 and 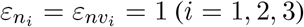. Further, let 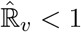. The equations for the infected compartments of this special case of the model can be re-written in terms of the next generation matrices (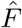 and 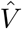) as follows:

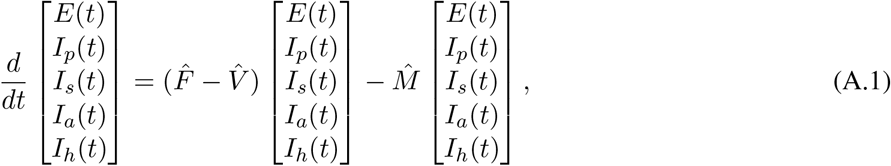

where (with *S*^*∗*^ and *A*^*∗*^ as defined in Section 3),

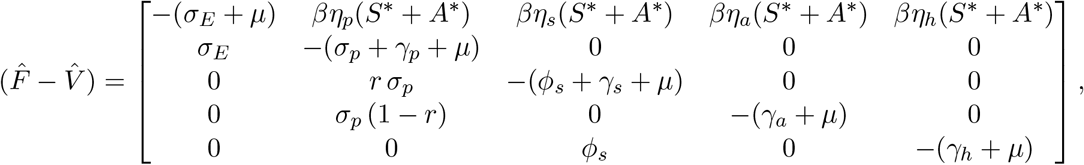

and,

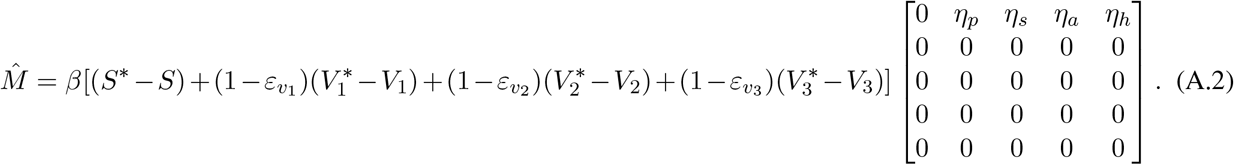

Since 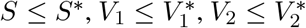 and 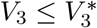 for all *t >* 0 in Ω_*∗∗*_, it follows that the matrix 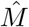, defined in (A.2), is non-negative. Hence, the equation (A.1) can be re-written in terms of the following inequality:

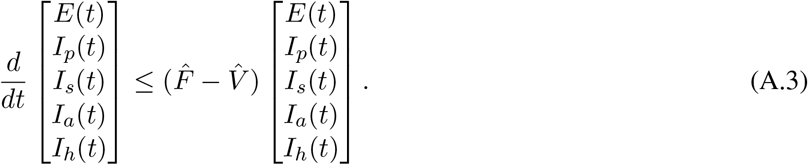

The proof is concluded the same way as in the proof of Theorem 3.2. Thus, the DFE (*E*_0_) of the special case of the model (2.1) (with *δ*_*p*_ = *δ*_*s*_ = *δ*_*a*_ = *δ*_*h*_ = 0 and 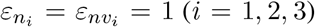) is globally-asymptotically stable in Ω_*∗∗*_ whenever 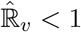.

## Notes

### Competing Interest Statement

The authors have declared no competing interest.

### Funding Statement

This study did not receive any funding

